# Platelet Activating Immune Complexes Identified in COVID-19 Associated Coagulopathy

**DOI:** 10.1101/2020.11.04.20226076

**Authors:** Ishac Nazy, Stefan D Jevtic, Jane C Moore, Angela Huynh, James W Smith, John G Kelton, Donald M Arnold

## Abstract

Thrombosis is a prominent feature of coronavirus disease 2019 (COVID-19) and often occurs in patients who are critically ill; however, the mechanism is unclear. This COVID-19 associated coagulopathy (CAC) shares features with heparin-induced thrombocytopenia (HIT), including mild thrombocytopenia and thrombosis. We thus tested 10 CAC patients for anti-PF4/heparin antibodies and functional platelet activation. HIT was excluded in all samples based on anti-PF4/heparin antibody and serotonin release assay results. Of note, 6 CAC patients demonstrated platelet activation by the serotonin release assay that was inhibited by FcγRIIA receptor blockade, confirming an IgG-specific immune complex (IC)-mediated reaction. Platelet activation was independent of heparin, but inhibitable by both therapeutic and high dose heparin. All 6 samples were positive for IgG-specific antibodies targeting the receptor binding domain (RBD) or the spike protein of the SARS-CoV-2 virus. These samples were additionally characterized by significant endothelial activation, shown by increased von Willebrand factor antigen and activity. ADAMTS13 activity was not severely reduced, and ADAMTS13 inhibitors were not present, ruling out thrombotic thrombocytopenic purpura. Our study thus identifies platelet-activating ICs as a mechanism that contributes to CAC thrombosis.

**Scientific Category:** Thrombosis and Hemostasis

**Key Points:**

- Patients with COVID-19 thrombosis have immune complexes that activate platelets through FcγRIIA signalling
- Patients with COVID-19 thrombosis demonstrate increased VWF antigen and activity that is not related to severe ADAMTS13 reduction

## Introduction

Coronavirus disease 2019 (COVID-19) is a highly transmissible viral infection caused by the severe acute respiratory syndrome coronavirus 2 (SARS-CoV-2) and has resulted in a global pandemic.^1,2^ Critically ill patients with COVID-19 can have prominent coagulation abnormalities, including mild thrombocytopenia and diffuse arterial and venous thrombosis.^3-6^ While the mechanism of COVID-19 associated coagulopathy (CAC) is unclear, this clinical presentation shares many features of heparin-induced thrombocytopenia (HIT), namely thrombocytopenia and thrombosis during critical illness.^7^

HIT is a prothrombotic disorder that typically presents as thrombocytopenia related to heparin treatment and is associated with a high risk of thrombosis. HIT is caused by IgG-specific antibodies targeting platelet-factor 4 (PF4) that form immune complexes (IC) and cause platelet activation through the FcγRIIA receptor.^8,9^ Recent reports have speculated that ICs also contribute to the pathobiology of severe COVID-19.^10^ In this report, we describe platelet-activating ICs in CAC patients. These ICs are not formed from IgG-specific anti-PF4/heparin antibodies and occur in combination with significant endothelial cell activation. Platelet-activating ICs may thus be an important mechanism of CAC thrombosis.

## Study Design and Methods

Blood samples from 10 critically ill patients with COVID-19 were referred to the McMaster Platelet Immunology Laboratory (MPIL) for HIT testing. Demographic data were obtained including sex, platelet nadir, heparin use, thrombotic event, admission diagnosis, and outcome. Control samples included 8 convalescent COVID-19 positive patients; 5 pre-pandemic HIT positive samples (HIT); and 7 pre-pandemic healthy controls (HC).

Testing for anti-PF4/heparin antibodies was done using an anti-PF4/heparin enzymatic immunoassay (EIA, LIFECODES PF4 enhanced assay, Immucor GTI Diagnostics, Waukesha, Wisconsin) for IgG, IgM, and IgA PF4-heparin antibodies. If positive, an in-house, IgG-specific anti-PF4/heparin EIA was performed.^11^ All samples were then tested for functional platelet activation in the serotonin release assay (SRA) with heparin (0.1, 0.3, and 100 U/mL) as previously described. An anti-human CD32 antibody (IV.3) was added to the SRA to confirm FcγRIIA engagement.^12^

Testing for IgG-, IgA- and IgM specific antibodies against the RBD and spike protein of SARS-CoV-2 virus was done using our in-house ELISA.^13^

As a measure of endothelial activation, VWF antigen levels were assessed by the HemosIL von Willebrand Factor antigen automated chemiluminescent immunoassay (Instrumentation Laboratory, Bedford, MA). VWF activity was measured using the Innovance VWF Ac Assay (OPHL03, Siemens, Marburg, Germany). ADAMTS13 metalloproteinase activity and anti-ADAMTS13 antibody were tested for all patients, as previously described, to determine whether VWF changes were related to ADAMTS13 activity.^14^ Data represent mean ± SEM and are analyzed by Student t test. Results are considered significant at *P* < 0.05.

This study was approved by the Hamilton Integrated Research Ethics Board.

## Results and Discussion

Blood samples were referred to the MPIL for HIT testing from 10 CAC patients (Supplementary Table 1). All patients developed thrombocytopenia and had been exposed to unfractionated heparin. All 5 patients for whom full clinical data were available had thrombosis; thrombosis data were unavailable for the remaining 5 patients. Initial testing for IgG, IgA, and IgM anti-PF4 antibodies was negative in 5 CAC samples (OD_405nm_ < 0.4), thus excluding HIT (Supplementary Table 2). IgG-specific anti-PF4 antibodies were then tested in the 5 positive CAC samples; 2 were negative (OD_405nm_ < 0.45), and 3 were weakly positive (OD_405nm_ range 0.495-0.931). Of these weakly positive samples, the positive predictive value of HIT is < 1.4% (Supplementary Table 2).^15^ All 10 CAC samples were then tested in the SRA where none demonstrated heparin-dependent platelet activation (Figure 1A). This combination of anti-PF4/heparin EIA results with heparin-independent platelet activation excludes HIT in all samples.^12^

**Figure 1:**
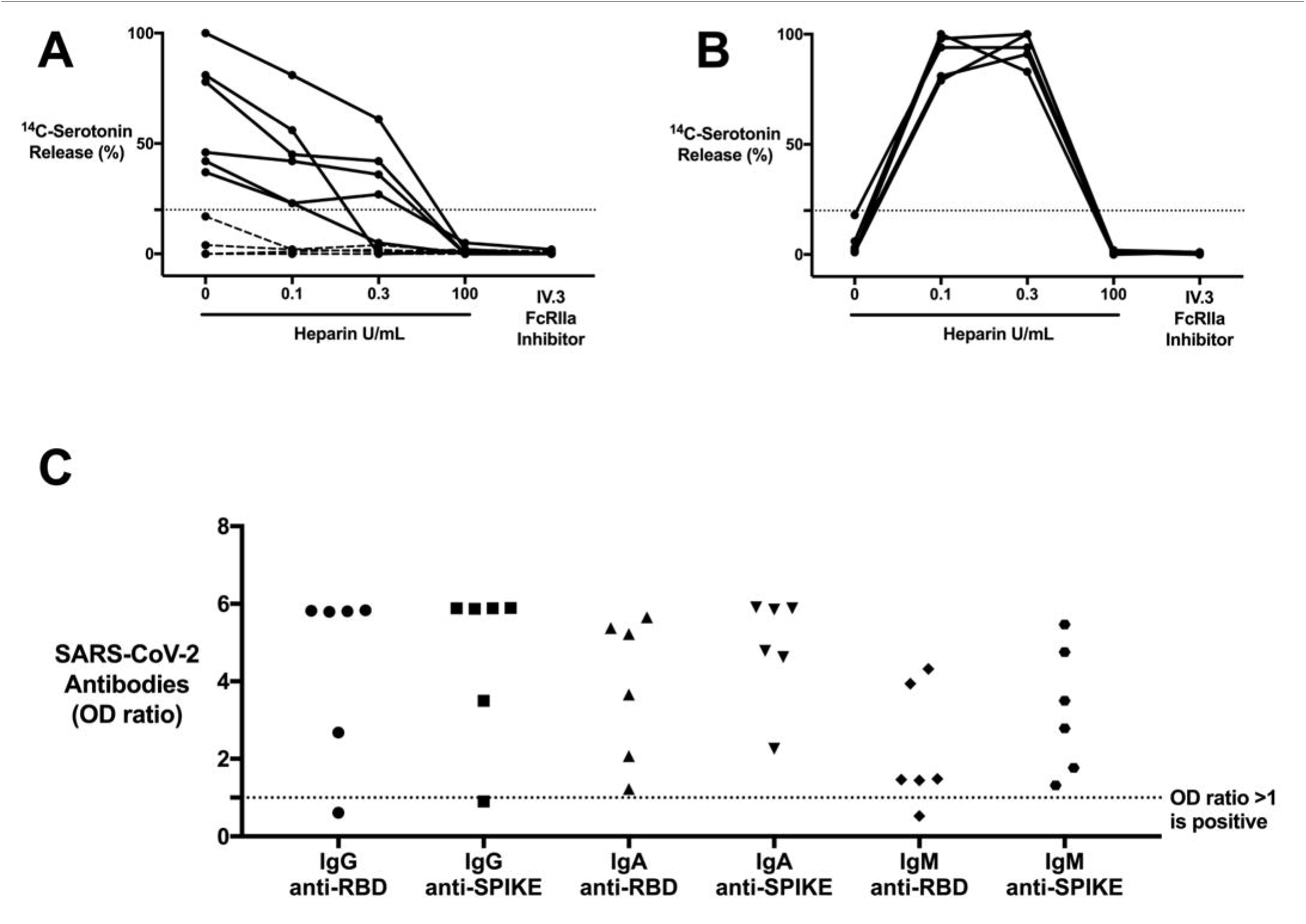
CAC patients with COVID-19 antibodies contain ICs that are capable of activating platelets in the SRA in a manner that is unique from HIT ICs. (A) CAC (n = 10) patient sera compared to (B) HIT patient (n = 5) sera, serving as a control, in the SRA. ^14^C-serotonin release was measured in the presence of increasing heparin doses or with addition of IV.3 (FcγRIIA inhibitor). ^14^C-serotonin release >20% is positive in the SRA (horizontal dashed line). Most CAC patient sera (n = 6, solid line) demonstrate heparin-independent platelet activation, as opposed to classic HIT controls. Platelet activation was inhibited with IV.3 in both groups. (C) IgG, IgA, and IgM COVID-19 antibodies in platelet-activating CAC patient sera (n = 6). Antibodies were measured in the SARS-CoV-2 ELISA and include RBD and spike protein specificity. Values are shown as a ratio of observed optical density to the determined assay cut-off optical density. Values above 1 ratio are considered positive in the SARS-CoV-2 ELISA.

6 CAC samples demonstrated significant platelet activation in the absence of heparin (Figure 1A). Platelet activation was inhibited with all heparin concentrations (0.1, 0.3 U and 100 U/mL), as opposed to the classic, heparin-dependent activation seen in HIT patients with therapeutic heparin levels (Figure 1A and 1B). Addition of IV.3 (FcγRIIA inhibitor) inhibited platelet activation, confirming an IgG-specific IC-mediated reaction (Figure 1A and 1B). Since certain virus-targeting, IgG-specific antibodies are known to form ICs, we tested for COVID-19 antibodies using our anti-SARS-CoV-2-ELISA.^13^ All 6 CAC samples contained IgG-specific antibodies against RBD, spike, or both proteins (Figure 1C), while none of the controls had anti-SARS-CoV-2 antibodies. Convalescent plasma from non-critically ill COVID-19 patients did not activate platelets in the SRA, despite having high titres of anti-SARS-CoV-2 antibodies, indicating that antibodies alone are insufficient for platelet activation (data not shown). A subset of CAC patient sera thus contain ICs that mediate platelet activation via FcγRIIA signalling.

ICs are known to trigger endothelial cell activation in diseases such as HIT and lupus vasculitis with subsequent VWF release.^16,17^ We therefore tested VWF antigen, which was markedly elevated in CAC samples (range, 2.1-10.1 U/mL; mean = 5.9 U/mL; normal range, 0.5-1.5 U/mL) compared with healthy controls (range 0.9-1.5 U/mL; mean = 1.2 U/mL) (Figure 2A). Ristocetin activity, as a marker of VWF activity, was also significantly increased (range, 2.2-8.52 U/mL; mean 4.6 U/mL; normal range 0.8-1.8 U/mL) compared to healthy controls (range, 0.53-1.67 U/mL, mean = 1.18 U/mL) (Figure 2B). These data confirm significant endothelial cell activation in CAC samples containing platelet-activating ICs.

**Figure 2:**
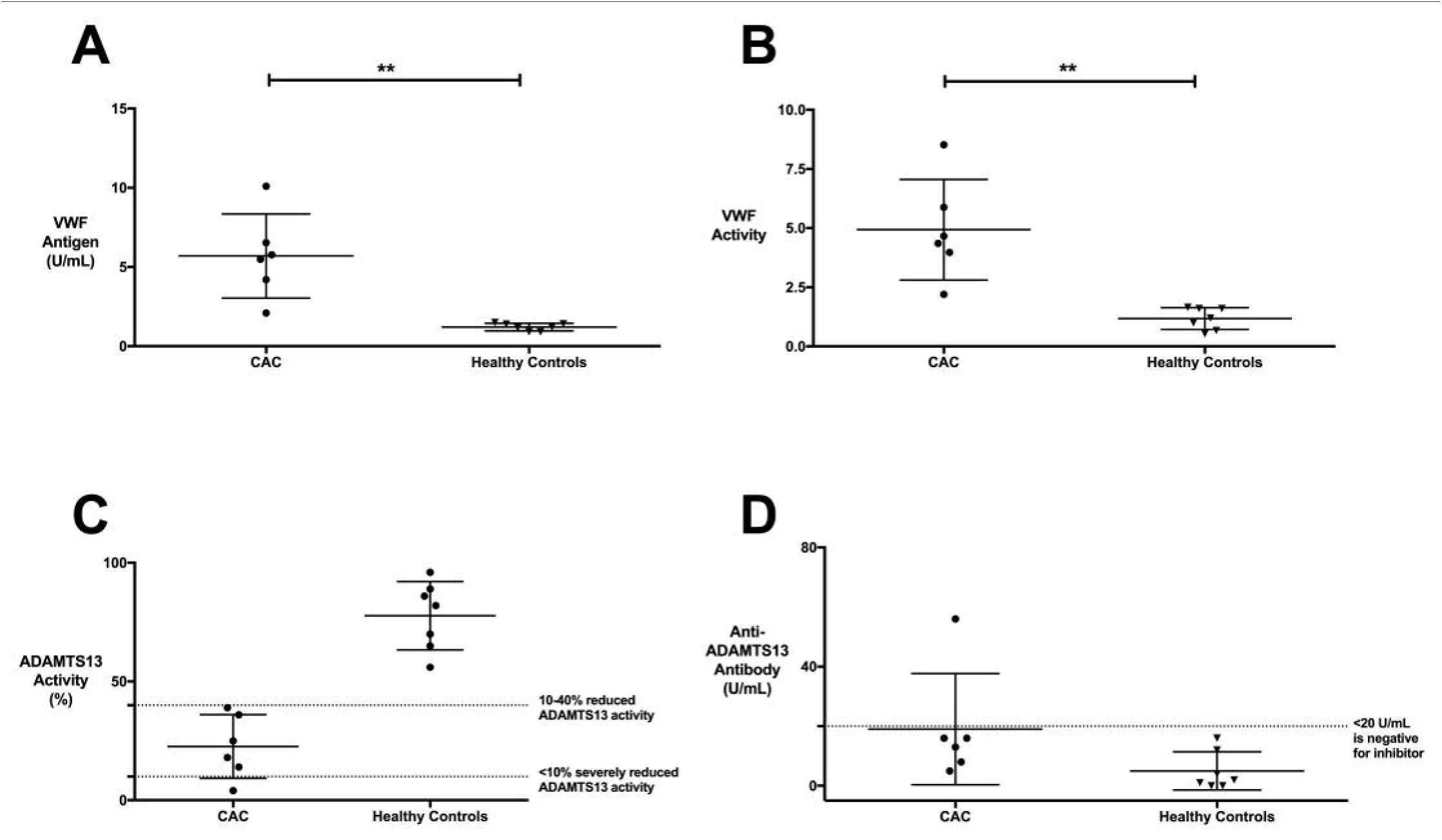
CAC samples with platelet-activating ICs show evidence of significant endothelial activation. (A) VWF antigen and (B) VWF activity are significantly elevated in CAC compared to healthy controls. (C) ADAMTS13 activity is moderately reduced (10-40%) in CAC as compared to healthy controls without an associated increase in (D) anti-ADAMTS13 antibody. One CAC sample was positive for presence of an anti-ADAMTS13 antibody, but this sample did not correspond to a severe reduction in ADAMTS13 activity. Therefore, none of the CAC samples meet criteria for TTP and are more in keeping with enhanced endothelial activation secondary to a secondary thrombotic microangiopathy. Data represent 6 CAC samples that demonstrated platelet-activating properties in the SRA. \*\**P* < 0.01, 2-tailed, unpaired Student *t*-test.

To determine if this VWF increase was related to ADAMTS13 metalloproteinase function, we tested for ADAMTS13 activity and the presence of anti-ADAMTS13 antibody. ADAMTS13 activity was moderately reduced (< 40%) in all CAC samples but only severely deficient (< 10%) in 1 (Figure 2C). Only 1 sample contained anti-ADAMTS13 antibody, which did not correspond to severe ADAMTS13 deficiency (Figure 2D). The elevated VWF is therefore not secondary to severe ADAMTS13 reduction.

We have described our findings in CAC samples obtained from COVID-19 patients with a clinical suspicion for HIT. Similar to HIT, certain patient samples contain ICs capable of mediating platelet activation. However, in contrast to HIT, CAC ICs are not formed of anti-PF4/heparin antibodies and are capable of platelet activation in the absence of heparin. The lack of anti-PF4/heparin antibodies and presence of heparin-independent SRA activation strongly rules out HIT. Furthermore, these platelet-activating ICs are inhibited by FcγRIIA blockade and heparin, including therapeutic (0.1 and 0.3 U/mL) and high (100 U/mL) doses. This is an important observation that highlights a novel mechanism for CAC ICs in promoting thrombosis.

We also confirm that endothelial cell activation contributes to coagulopathy, as previously shown.^18^ We demonstrate that this is not secondary to severe ADAMTS13 activity reduction (<10 %) or the presence of anti-ADAMTS13 antibody, as described in thrombotic thrombocytopenic purpura (TTP). This pattern is more consistent with a secondary thrombotic microangiopathy and is likely compounded by the presence of platelet-activating ICs.^19,20^

A limitation of this study is the inability to characterize the specificity of these platelet-activating ICs. It is possible that the ICs are composed of COVID-19 virus-antibody complexes, as seen with H1N1 viral infection.^21^ This is further supported by inhibition with therapeutic heparin, since heparin binds the SARS-CoV-2 RBD to cause a conformational change with altered binding specificity, potentially disrupting the ICs and inhibiting platelet activation.^22^ Regardless, the data presented here clearly outline the characteristics of these ICs and differentiate them from other severe coagulation disorders, including HIT and TTP.

We thus propose a model whereby certain severe CAC patients feature a novel IC-mediated thrombotic microangiopathy that is characterized by significant platelet and endothelial cell activation. These ICs can produce a highly prothrombotic state resembling HIT but with unique platelet activating properties.

## Data Availability

Additional data can be obtained by emailing the corresponding author.

## Acknowledgements

Funding support for this work was provided by a grant from the Ontario Research Fund (ORF #2426729), COVID-19 Rapid Research Fund awarded to Dr. Ishac Nazy (#VR2-173204), and Academic Health Sciences Organization (HAHSO) grant awarded to Dr. Donald M Arnold (#HAH-21-02).

## Authorship Contributions

IN designed the research, analyzed and interpreted data and wrote the manuscript. SJ, JCM, AH and JWS carried out the described studies, analyzed data, and wrote the manuscript. JGK and DMA designed the research and wrote the manuscript. All authors reviewed and approved the final version of the manuscript.

