## Supplementary Materials for "Platelet Activating Immune Complexes Identified in COVID-19 Associated Coagulopathy"

**Supplementary Table 1: Critically Ill COVID-19 Coagulopathy Patient Characteristics**

| **Sample ID (CAC)** | **Sex** | **Heparin Use** | **Platelet Nadir (10^6^/L)** | **Thrombosis** | **Outcome** |
| --- | --- | --- | --- | --- | --- |
| 1 | M | UFH | 69,000 | Present | Discharged |
| 2 | M | UFH | N/A | N/A | Deceased |
| 3 | M | UFH | N/A | N/A | N/A |
| 4 | M | UFH | 58,000 | Present | Deceased |
| 5 | M | UFH | 12,000 | N/A | Deceased |
| 6 | M | UFH | 96,000 | N/A | Deceased |
| 7 | M | UFH | 61,000 | Present | Deceased |
| 8 | M | UFH | 42,000 | Present | Discharged |
| 9 | F | UFH | 11,000 | N/A | N/A |
| 10 | F | UFH | 72,000 | Present | Deceased |

N/A = not available.

UFH = unfractionated heparin

**Supplementary Table 2: Anti-PF4/Heparin IgG, IgA, and IgM Antibody Levels in CAC Samples Detected by EIA.**

| **Sample ID (CAC)** | **IgG, IgM, IgA (OD_405nm_)*** | **IgG-specific (OD_405nm_)**** | **Positive Predictive Value of HIT (%)** |
| --- | --- | --- | --- |
| 1 | 0.506 | 0.235 | < 0.5 |
| 2 | 0.102 | N/A |  |
| 3 | 0.867 | 0.495 | 1.4 |
| 4 | 0.168 | N/A |  |
| 5 | 3.155 | 0.583 | 1.4 |
| 6 | 0.456 | 0.103 | < 0.5 |
| 7 | 1.64 | 0.931 | 1.4 |
| 8 | 0.086 | N/A |  |
| 9 | 0.261 | N/A |  |
| 10 | 0.049 | N/A |  |

Samples that were initially positive in the IgG, IgM, IgA anti-PF4/heparin EIA were subsequently tested in the IgG-specific anti-PF4/heparin EIA. The positive predictive value of HIT is based on the IgG-specific anti-PF4/heparin EIA result as previously described.

N/A = not available.

* Positive OD_405nm_ > 0.4 in the IgG, IgA, IgM anti-PF4/heparin EIA

** Positive OD_405nm_ > 0.45 in the IgG-specific anti-PF4/heparin EIA
